# Knowledge towards breast cancer, and breast self-examination practices and its barriers among university female students in Bangladesh: Findings from a cross-sectional study

**DOI:** 10.1101/2021.10.20.21265262

**Authors:** Rumpa Sarker, Md. Saiful Islam, Mst. Sabrina Moonajilin, Mahmudur Rahman, Hailay Abrha Gesesew, Paul R Ward

## Abstract

Early diagnosis of breast cancer is the best approach towards its control that may result in alleviating related mortality and morbidity. This study aimed to evaluate knowledge about breast cancer and both practices and perceived barriers to breast self-examination among female university students in Bangladesh. A cross-sectional study was carried out with 400 female students of Jahangirnagar University, Bangladesh. Participants were sampled from female dormitories at the university from January to April 2020. Proportionate stratified random sampling was conducted to calculate the study sample from each dormitory. A pre-tested, semi-structured self-reported questionnaire was employed to collect data from participants during the survey periods. The questionnaire consists of demographic variables, items about knowledge about breast cancer, breast self-examination practices and its barriers. We applied descriptive and inferential statistics, and data were analyzed using SPSS. Participants aged between 18-26 years and comprised university students of 1^st^ year (20%), 2^nd^ year (24%), 3^rd^ year (22%), 4^th^ year (21%) and Master’s (14.%). 18% of them had reported positive family history (mother, aunt, sister/cousin, grandmother) of breast cancer. The overall mean score of total knowledge items was 15 (SD = 3) out of 43, with an overall correct rate of 34%. The mean score of total knowledge items was significantly higher (*p*<0.001) among Master’s students and students with family members who have breast cancer. Only one in five students (21%) ever practiced breast self-examination. The mean score of practice towards breast self-examination was significantly higher (*p*<0.001) among participants who reported having family member of breast cancer. It is noted that the total knowledge score about breast cancer and practice towards breast self-examination are significantly correlated with each other (*r* = 0.54; *p*<0.001). About 33% participants had reported that ‘lack of knowledge as the main barrier to practicing breast self-examination followed by ‘I do not have the symptoms’ (22%), and shyness/ uncomfortable feelings (17%). The study revealed low levels of knowledge about breast cancer and low breast self-examination practices. This implies the need of promotion and implementation of educational interventions programs that are social and culturally appropriate and suitable for specific age groups.

## Introduction

Breast cancer is a major global health concern and a prominent reason of mortality among females. It is the most frequent cancer among women globally, impacting 2.1 million women each year, and is predicted to grow to approximately 3.2 million new cases per year by 2050 [1]. There were 13,028 new breast cancer cases (19% of all cancer types among females) in Bangladesh in 2020 [2]. Although the incidence rate is higher among women older than 50 years, the rate of diagnosis with breast cancer among young women under 50 years of age have increased. Breast cancer also causes the greatest number of cancer-related deaths among women [3]. For example, an estimated 627,000 women died from breast cancer in 2018 in Bangladesh, contributing to nearly 15% of all cancer deaths among women [4]. Delay in diagnosis and seeking proper treatment from the primary symptomatic detection of breast cancer lowers the level of successful treatment outcome and thus decreased survival length [5]. But most breast cancer patients are diagnosed in developing countries, including Bangladesh, at an advanced stage due to a lack of understanding and insufficient access to health care facilities [6]. In Bangladesh, breast cancer is ranked as the 2^nd^ leading cancer after cervical cancer and together breast and cervical cancer account for 38% of all cancer among women.

The American Cancer Society recommends that women should be familiar with how their breasts normally feel via breast self-examination (BSE) and report any breast changes promptly to their health care providers. The ‘National Cancer Control Strategy and Plan of Action 2009-15’ in Bangladesh advocates encouraging clinical breast examination (CBE) and BSE as breast cancer’s early detection approach for disease downstage and survival improvement [7]. In addition, the Breast Health Global Initiative (BHGI) guideline for low- and middle-income countries suggests BSE as the first step in preventing breast cancer [8].

BSE is an easy, expedient, non-invasive and no-cost way to check out women’s own breasts to find any changes in their breasts that can be an early symptom of breast cancer in the initial phase when the condition can be treated with successful outcome and thus increasing survival rate from breast cancer. BSE aids women by making them conversant about how their breasts should look and feel thus leading to ‘breast awareness’ and also enable them to identify changes in their breasts in the initial stage [9]. It can be performed on a regular basis, at any age and is suitable for low resource countries like Bangladesh. Conversely, mammography screening is not a practical approach to pursue breast cancer prevention due to its high costs for the health system and individual women (in terms of out-of-pocket costs). Although inappropriate or inaccurate BSE enactment may produce both false positives and false negatives for women, BSE is still regarded as a legitimate and realistic alternative for early breast cancer screening in women [10].

In Bangladesh, several studies have reported poor knowledge, and awareness of breast cancer and lack of adherence to any recommended screening method of breast cancer, including BSE [11]. However, very few studies have been conducted with young and educated females who are within their reproductive age. It is crucial to assess their level of knowledge and practice of BSE as this assessment of knowledge may reflect the awareness level of a large proportion of the population. Although breast cancer incidence is lower in this age group of women than for older women, it is important for younger women to practice BSE in order to identify changes in breast tissue in the future and hopefully prevent incurable, late-stage cancers. Thus, using the case of Jahangirnagar University, this study aimed to investigate the knowledge of symptoms, risk factors, treatment modalities and screening methods of breast cancer among young females who represent the most educated segment of population, as well as to examine the practice of BSE and the barriers that are hindering the practice of BSE.

## Methodology

### Participants and procedures

A cross-sectional study was carried out with 400 female students who enrolled in Bachelor and Master’s programs of Jahangirnagar University (fully residential university of Bangladesh). The study was solely conducted in the 8 female dormitories and data were collected over four months from January-April 2020. At first, for sample size determination, Yamane’s simplified sampling formula was used. By that, we got 386 as sample size and for our convenience, we took 400 respondents. After that, the proportionate stratified random sampling was conducted based on the population proportion of dormitories to calculate the study sample from each dormitory. In this approach, each stratum sample size is directly proportional to the population size of the entire population of strata.

The inclusion criteria included: (i) being 18 years of above; (ii) being female university students who reside at dormitories; and (iii) being willing to take part in the survey. The exclusion criteria were being below 18 years old and having incomplete responses.

### Study instruments

A pre-tested, self-reported, semi-structured questionnaire including informed consent, socio-demographic information and questions related to knowledge towards breast cancer, BSE practices and its barriers, was prepared for the study through extensive literature review [12-16]. In order to add further face validity, the questionnaire was reviewed by an external reviewer who is an oncologist with extensive experience of consulting women in Bangladesh about breast cancer prevention, diagnosis and prognosis. Likewise, a pilot test was conducted to assess the readability of the questionnaire. The questionnaire was finalized after incorporating minor amendments based on the participants’ feedback during the piloting periods. The survey questionnaire is attached in the **supplementary file 1**.

#### Socio-demographic information

Socio-demographic information was recorded during the survey including age, study year (1^st^/ 2^nd^/ 3^rd^/ 4^th^/ Master’s), marital status (unmarried/ unmarried), family history of breast cancer (yes/ no), and relationship with breast cancer affected patient (mother/ sister/ cousin/ aunt/ grandmother).

#### Knowledge towards breast cancer measures

To assess the participants’ knowledge of breast cancer, a total of 43 questions regarding breast cancer (i.e., 8 for symptoms, 10 for risk factors, 6 for treatment, 8 for prevention, 5 for screening, and 5 for process of BSE) were asked during the survey. Each question has three possible responses (i.e., yes/no/don’t know). The distributions (frequencies and percentages) of all questions are presented in **supplementary file 2**.

#### BSE practices and its barriers measures

A single construct (i.e., *Have you ever self-examined your breast for breast cancer?*) was used to assess the BSE with binary responses (yes/no). In addition, the barriers of BSE were also recorded.

### Data analysis

Data analysis was performed using the SPSS version 25. Descriptive statistics such as frequencies and percentages were computed for categorical variables; whereas, means and standard deviations were for continuous variables. Some first-order analyses (i.e., t-tests and one-way ANOVA) were performed to assess the association between independent and dependent variables. The Pearson correlation test was also carried out to find out the correlations between two continuous variables. The significance level (*p*-value) was set at 0.05.

### Ethical considerations

The study protocol was reviewed and approved by the local Intuitional Review Board of Jahangirnagar University in Bangladesh. Before starting data collection, participants were informed about the objectives, methodology and the approximate time to complete the survey. Then written informed consent was obtained from each participant. Likewise, participants were also assured that all information related to them will be kept confidential.

## Results

A total of 400 female participants aged between 18-26 years were included in the final analysis. The participants comprised of university students of 1^st^ year (19.5%), 2^nd^ year (23.5%), 3^rd^ year (21.8%), 4^th^ year (21.0%) and Master’s (14.2%) (Table 1). Most of them were unmarried (86.0%). A sizable minority reported they had family history of breast cancer (18.3%). The participants also reported the relationship with affected people as follows: mother (11.6%; n = 8), sister/cousin (24.6%; n = 17), aunt (40.6%; n = 28), and grandmother (23.2%; n = 16).

**Table 1.** Characteristics of participants.

| Variables | n | (%) |
| --- | --- | --- |
| **Age** |  |  |
| 18-20 | 141 | (35.3) |
| 21-23 | 124 | (31.0) |
| 24-26 | 135 | (33.8) |
| **Study year** |  |  |
| 1st | 78 | (19.5) |
| 2nd | 94 | (23.5) |
| 3rd | 87 | (21.8) |
| 4th | 84 | (21.0) |
| Master's | 57 | (14.2) |
| **Marital status** |  |  |
| Unmarried | 344 | (86.0) |
| Married | 56 | (14.0) |
| **Family history of breast cancer** |  |  |
| Yes | 73 | (18.3) |
| No | 327 | (81.8) |
| **Relationship with affected patient** |  |  |
| Mother | 8 | (11.6) |
| Sister/cousin | 17 | (24.6) |
| Aunt | 28 | (40.6) |
| Grandmother | 16 | (23.2) |

### Knowledge about breast cancer’s symptoms, risk, treatment, prevention, screening, and process of screening of breast

The mean score of knowledge about breast cancer’s symptoms was 2.94 (SD = 1.14) out of 8, with an overall correct rate of 36.8% (Table 2). The most common symptoms that were addressed frequently were ‘color change of breast including redness or flaky skin’ (48.3%), ‘new lump in the breast or armpit’ (42.8%), ‘changes in breast shape and size including inverted nipple’ (40.3%), ‘nipple discharge other than breast milk including blood or pus’ (29.8%). The mean score of knowledge about breast cancer symptoms was significantly higher among participants of 5^th^ year (Master’s). The mean score of knowledge about breast cancer’s risk was 3.35 (SD = 1.19) out of 10, with an overall correct rate of 33.5%. The mean score of knowledge about breast cancer’s risk was significantly higher among participants who reported being 24-26 years, Master’s student, and having other sources of information. The mean score of knowledge about breast cancer treatment was 1.80 (SD = 0.93) out of 6, with an overall correct rate of 30.0%. The mean score of knowledge about breast cancer treatment was significantly higher among participants who reported being 24-26 years, Master’s student, and having other sources of information.

**Table 2.** Distribution of participants’ knowledge about breast cancer’s symptoms, risk, treatment, prevention, screening, and process of breast self-examination.

| Variables | Symptoms |  |  | Risk |  |  | Treatment |  |  | Prevention |  |  | Screening |  |  | Process of BSE |  |  |
| --- | --- | --- | --- | --- | --- | --- | --- | --- | --- | --- | --- | --- | --- | --- | --- | --- | --- | --- |
|  | Total score=8 |  |  | Total score=10 |  |  | Total score=6 |  |  | Total score=8 |  |  | Total score=5 |  |  | Total score=5 |  |  |
|  | Mean (SD) | t/F | p-value | Mean (SD) | t/F | p-value | Mean (SD) | t/F | p-value | Mean (SD) | t/F | p-value | Mean (SD) | t/F | p-value | Mean (SD) | t/F | p-value |
| **Age** |  |  |  |  |  |  |  |  |  |  |  |  |  |  |  |  |  |  |
| 18-20 | 2.82 (1.29) | 1.81 | 0.166 | 3.03 (1.16) | 8.37 | **<0.001** | 1.89 (1.02) | 4.77 | **0.009** | 3.34 (1.11) | 1.23 | 0.294 | 1.76 (0.54) | 1.42 | 0.243 | 1.56 (1.9) | 0.26 | 0.772 |
| 21-23 | 3.09 (1.04) |  |  | 3.48 (1.23) |  |  | 1.59 (0.96) |  |  | 3.13 (1.12) |  |  | 1.75 (0.47) |  |  | 1.48 (1.8) |  |  |
| 24-26 | 2.93 (1.05) |  |  | 3.56 (1.12) |  |  | 1.9 (0.75) |  |  | 3.3 (1.2) |  |  | 1.84 (0.61) |  |  | 1.64 (1.89) |  |  |
| **Study year** |  |  |  |  |  |  |  |  |  |  |  |  |  |  |  |  |  |  |
| 1<sup>st</sup> year | 2.55 (1.33) | 6.40 | **<0.001** | 3.22 (1.12) | 5.84 | **0.016** | 1.86 (0.81) | 4.50 | **0.001** | 3.14 (1.2) | 2.65 | **0.033** | 1.86 (0.55) | 0.66 | 0.623 | 1.79 (1.98) | 0.82 | 0.516 |
| 2<sup>nd</sup> year | 2.83 (1.08) |  |  | 2.98 (1.24) |  |  | 1.62 (1.05) |  |  | 3.03 (1.14) |  |  | 1.79 (0.53) |  |  | 1.52 (1.89) |  |  |
| 3<sup>rd</sup> year | 2.98 (1.04) |  |  | 3.26 (1.19) |  |  | 1.63 (0.88) |  |  | 3.3 (1.06) |  |  | 1.78 (0.49) |  |  | 1.55 (1.74) |  |  |
| 4<sup>th</sup> year | 2.93 (1.07) |  |  | 3.61 (0.98) |  |  | 1.82 (0.78) |  |  | 3.26 (1.23) |  |  | 1.77 (0.6) |  |  | 1.32 (1.74) |  |  |
| 5<sup>th</sup> year (Master's) | 3.4 (1.01) |  |  | 3.68 (1.27) |  |  | 2.14 (0.96) |  |  | 3.58 (1.05) |  |  | 1.88 (0.59) |  |  | 1.85 (2) |  |  |
| **Marital status** |  |  |  |  |  |  |  |  |  |  |  |  |  |  |  |  |  |  |
| Unmarried | 2.92 (1.16) | 1.36 | 0.245 | 3.36 (1.18) | 0.19 | 0.664 | 1.82 (0.94) | 1.48 | 0.225 | 3.28 (1.14) | 1.16 | 0.282 | 1.83 (0.54) | 2.32 | 0.128 | 1.58 (1.84) | 0.06 | 0.813 |
| Married | 3.11 (1.04) |  |  | 3.29 (1.25) |  |  | 1.66 (0.79) |  |  | 3.11 (1.19) |  |  | 1.71 (0.56) |  |  | 1.52 (1.98) |  |  |
| **Family member of breast cancer** |  |  |  |  |  |  |  |  |  |  |  |  |  |  |  |  |  |  |
| Yes | 3.08 (1.2) | 1.34 | 0.248 | 3.48 (1.24) | 1.05 | 0.305 | 1.88 (0.96) | 0.61 | 0.434 | 3.18 (1.22) | 0.46 | 0.500 | 1.73 (0.65) | 2.50 | 0.115 | 4.25 (1.7) | 341.87 | **<0.001** |
| No | 2.91 (1.13) |  |  | 3.32 (1.18) |  |  | 1.78 (0.92) |  |  | 3.28 (1.13) |  |  | 1.84 (0.52) |  |  | 0.98 (1.28) |  |  |
| **Source of information** |  |  |  |  |  |  |  |  |  |  |  |  |  |  |  |  |  |  |
| Friends/relatives | 2.92 (1.21) | 1.20 | 0.311 | 3.09 (1.27) | 4.40 | **0.002** | 1.77 (1.04) | 0.20 | 0.941 | 3.19 (1.2) | 1.46 | 0.214 | 1.77 (0.55) | 0.63 | 0.644 | 1.84 (1.99) | 1.64 | 0.163 |
| Social media | 2.86 (1.2) |  |  | 3.08 (1.19) |  |  | 1.8 (0.89) |  |  | 3.38 (1.25) |  |  | 1.81 (0.53) |  |  | 1.27 (1.74) |  |  |
| Health campaign | 2.84 (1.1) |  |  | 3.55 (1.13) |  |  | 1.86 (0.92) |  |  | 3.22 (1.08) |  |  | 1.86 (0.6) |  |  | 1.65 (1.93) |  |  |
| Doctors | 3.18 (1.03) |  |  | 3.47 (1.22) |  |  | 1.74 (0.9) |  |  | 3.47 (1.2) |  |  | 1.87 (0.46) |  |  | 1.27 (1.57) |  |  |
| Others | 3.09 (1.1) |  |  | 3.73 (0.91) |  |  | 1.8 (0.79) |  |  | 3 (0.85) |  |  | 1.76 (0.57) |  |  | 1.78 (1.94) |  |  |
| **Practice of BSE** |  |  |  |  |  |  |  |  |  |  |  |  |  |  |  |  |  |  |
| Yes | 2.96 (1.16) | 0.04 | .840 | 3.29 (1.29) | 0.24 | .627 | 1.85 (0.93) | 0.28 | .598 | 3.13 (1.26) | 1.41 | .236 | 1.82 (0.6) | 0.01 | .909 | 4.98 (0.15) | 3792.90 | **<0.001** |
| No | 2.94 (1.14) |  |  | 3.37 (1.17) |  |  | 1.79 (0.93) |  |  | 3.3 (1.11) |  |  | 1.82 (0.53) |  |  | 0.65 (0.64) |  |  |

The mean score of knowledge about breast cancer prevention was 3.26 (SD = 1.14) out of 8, with an overall correct rate of 36.2%. The mean score of knowledge about breast cancer prevention was significantly higher among participants who reported being Master’s student. The mean score of knowledge about breast cancer screening was 1.82 (SD = 0.55) out of 5, with an overall correct rate of 36.4%. The mean score of knowledge about breast cancer screening was not significantly differ among participants in terms of socio-demographic and source of information. The mean score of knowledge about BSE process was 1.57 (SD = 1.86) out of 5, with an overall correct rate of 31.4%. The mean score of knowledge about BSE process was significantly higher among participants who reported having family members of breast cancer and those who have had practiced BSE ever.

Overall, the mean score of total knowledge items was 14.74 (SD = 3.15) out of 43, with an overall correct rate of 34.3% (Table 3). The mean score of total knowledge items was higher among students of Master’s and having family members of breast cancer. The participants’ sources of information about breast cancer are presented in Figure 1.

**Table 3.** Participants’ total knowledge and practice.

|  | Knowledge |  |  | Practice |  |  |
| --- | --- | --- | --- | --- | --- | --- |
|  | Mean (SD) | t/F | p-value | Mean (SD) | t/F | p-value |
| **Age** |  |  |  |  |  |  |
| 18-20 | 14.54 (3.25) | 1.83 | 0.162 | 0.22 (0.42) | 0.19 | 0.825 |
| 21-23 | 14.52 (3.16) |  |  | 0.19 (0.4) |  |  |
| 24-26 | 15.16 (3.01) |  |  | 0.22 (0.42) |  |  |
| **Study year** |  |  |  |  |  |  |
| 1<sup>st</sup> year | 15.14 (3.19) | 3.47 | **0.008** | 0.26 (0.44) | 0.81 | 0.520 |
| 2<sup>nd</sup> year | 13.77 (3.34) |  |  | 0.21 (0.41) |  |  |
| 3<sup>rd</sup> year | 14.77 (3.01) |  |  | 0.18 (0.39) |  |  |
| 4<sup>th</sup> year | 14.82 (2.79) |  |  | 0.17 (0.37) |  |  |
| 5<sup>th</sup> year (Master's) | 15.34 (3.12) |  |  | 0.26 (0.44) |  |  |
| **Marital status** |  |  |  |  |  |  |
| Unmarried | 14.8 (3.2) | 0.80 | 0.371 | 0.21 (0.41) | 0.15 | 0.699 |
| Married | 14.39 (2.81) |  |  | 0.23 (0.43) |  |  |

| Family member of breast cancer |  |  |  |  |  |  |
| --- | --- | --- | --- | --- | --- | --- |
| Yes | 17.59 (2.5) | 89.24 | <0.001 | 0.84 (0.37) | 427.67 | <0.001 |
| No | 14.11 (2.92) |  |  | 0.07 (0.26) |  |  |
| Source of information |  |  |  |  |  |  |
| Friends/relatives | 14.58 (3.27) | 1.13 | 0.341 | 0.27 (0.45) | 1.52 | 0.195 |
| Social media | 14.2 (3.02) |  |  | 0.17 (0.37) |  |  |
| Health campaign | 14.97 (3.53) |  |  | 0.23 (0.42) |  |  |
| Doctors | 15 (2.37) |  |  | 0.13 (0.34) |  |  |
| Others | 15.16 (3.01) |  |  | 0.24 (0.43) |  |  |

**Figure 1.**
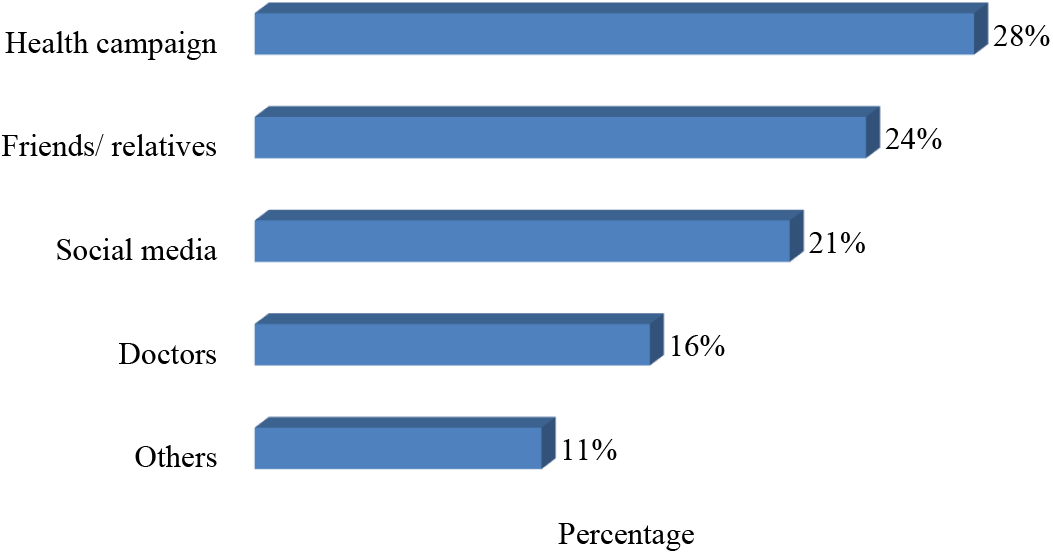
Source of information about breast cancer.

### Practices of breast self-examination

About one-fifth participants reported they had ever practiced BSE (21.3%). The mean score of practice towards BSE was significantly higher among participants who reported having family members of breast cancer. Descriptive statistics, and correlations between all outcome variables (i.e., knowledge about breast cancer’s symptoms, risk, treatment, prevention, screening, and process of breast self-examination, practice of breast self-examination) are presented in Table 4. It is noted that the total knowledge score about breast cancer and practice towards BSE are significantly correlated with each other (*r* = 0.54; *p*<0.001).

**Table 4.**
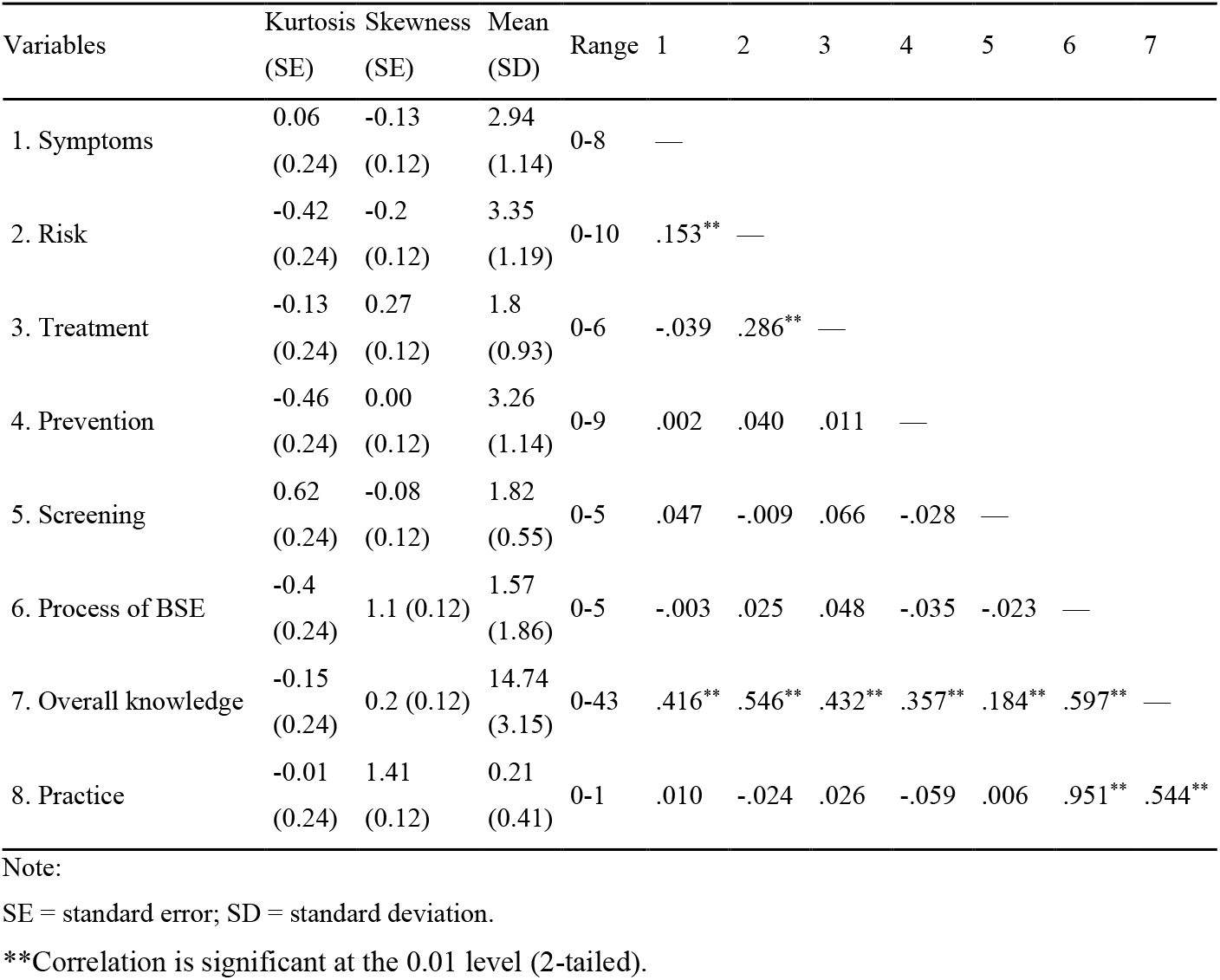
Descriptive statistics, and correlations between all outcome variables.

| Variables | Kurtosis<br>(SE) | Skewness<br>(SE) | Mean<br>(SD) | Range | 1 | 2 | 3 | 4 | 5 | 6 | 7 |
| --- | --- | --- | --- | --- | --- | --- | --- | --- | --- | --- | --- |
| 1. Symptoms | 0.06<br>(0.24) | -0.13<br>(0.12) | 2.94<br>(1.14) | 0-8 | — |  |  |  |  |  |  |
| 2. Risk | -0.42<br>(0.24) | -0.2<br>(0.12) | 3.35<br>(1.19) | 0-10 | .153** | — |  |  |  |  |  |
| 3. Treatment | -0.13<br>(0.24) | 0.27<br>(0.12) | 1.8<br>(0.93) | 0-6 | -.039 | .286** | — |  |  |  |  |
| 4. Prevention | -0.46<br>(0.24) | 0.00<br>(0.12) | 3.26<br>(1.14) | 0-9 | .002 | .040 | .011 | — |  |  |  |
| 5. Screening | 0.62<br>(0.24) | -0.08<br>(0.12) | 1.82<br>(0.55) | 0-5 | .047 | -.009 | .066 | -.028 | — |  |  |
| 6. Process of BSE | -0.4<br>(0.24) | 1.1 (0.12) | 1.57<br>(1.86) | 0-5 | -.003 | .025 | .048 | -.035 | -.023 | — |  |
| 7. Overall knowledge | -0.15<br>(0.24) | 0.2 (0.12) | 14.74<br>(3.15) | 0-43 | .416** | .546** | .432** | .357** | .184** | .597** | — |
| 8. Practice | -0.01<br>(0.24) | 1.41<br>(0.12) | 0.21<br>(0.41) | 0-1 | .010 | -.024 | .026 | -.059 | .006 | .951** | .544** |

Note:
SE = standard error; SD = standard deviation.
\*\*Correlation is significant at the 0.01 level (2-tailed).

### Barrier towards breast self-examination

About 33.3% of the participants have addressed ‘lack of knowledge’ as the main barrier to practicing BSE followed by ‘I do not have the symptoms’ (21.8%), ‘ shyness/ uncomfortable feelings’ (16.5%), ‘I don’t think it’s important (9.5%), ‘I know I will never have breast cancer’ (6.8%), ‘fear or being diagnosed of cancer’ (6.3%).

## Discussion

The present study explored the knowledge of breast cancer and practice of BSE among university students in Bangladesh. The present findings on the common symptoms are in consistent with a study in Saudi Arabia which also showed the moderate knowledge of ‘presence of lump in the breast’ (48%) and ‘ change in the shape of the breast or nipple’ (42.9%), although ‘inverted nipple’ was known as a warning sign of breast cancer by only 29% of the students [12]. In another study on female university students of Egypt, breast lump was the most commonly identified (81.6%) symptom of breast cancer [13], though the percentage was significantly higher than our study implying that the awareness regarding symptoms of breast cancer needs to be improved in Bangladesh.

Our findings on levels of knowledge about risk factors of breast cancer were also consistent with the Saudi Arabian study which also showed that only 42.3% of the study sample knew that lack of physical exercise increases the risk of breast cancer [12]. Consistent with our study, a number of studies with different groups of women in Malaysia have also found that a lack of knowledge of breast cancer was the most commonly identified addressed barrier to breast cancer prevention practices [18, 31]. A moderate number of respondents in our study had positive attitudes toward the early detection breast cancer, which has also been found in Iraq [19]. In the study conducted in Iraq, the participants had low knowledge on the treatment options for breast cancer - only 40.3% stated chemotherapy, 37.3% stated surgery, and 27.8% stated hormonal therapy. In comparison, a study conducted with female university students in Egypt found a significantly higher percentage of students recognized chemotherapy and surgery as treatment options [15], suggesting that students in Bangladesh may require further information on treatment options for breast cancer so they know it can be treated and thus are aware of the reasons and benefits for BSE. Interestingly, respondents in our study believed that alternative medicines (17.0%) and herbal treatments (20.5%) are effective treatment options for breast cancer. We need to kept in mind that our sample was women currently at university who one may assume have access to the most up-to-date information sources and are thus among the most literate, health literate and digitally literate women in Bangladesh. Therefore, if levels of knowledge are low in this group, we assume they would be even lower in other groups of women who do not go to university. As such, evidence-based educational materials may be required to provide women (at different literacy levels and therefore in different ways) in Bangladesh for effective treatments for breast cancer.

BSE was known to only 42.0% of the respondents in our study, a finding consistent with a study in Pakistan which found that only 40.3% of students knew about BSE [20]. CBE was the least known method for breast screening in our study, whereas 64.2% had heard of CBE in a different study conducted among women aged 30–59 years was conducted in Bangladesh in Bangladesh [21]. Also, in contrast with our study, a study with female university students in Egypt found that 74.2% know about BSE, 52.1% about mammogram and 48.3% about ultrasound [15], which is considerably higher than in our study. In our study, although 42.0% knew about BSE only 21.3% of the respondents had ever practiced BSE, which is similar to another study in Bangladesh whereby 46.7% of respondents knew about BSE but only 16.3% had practiced BSE ever [12]. Indeed, the practice rate of BSE by female students is low in a number of countries, such as Cameroon (38.5%), Yemen (17.4%), Iraq (19.7%), Turkey (20.3%), Egypt (6.1%), and Korea (27%) [15, 17, 19, 22-24].

In our study, there is a significant relationship between the overall knowledge score and practice of BSE implying that the more knowledgeable the participants are greater likely to practice breast self-examination. We are acutely aware that correlation does not equal causation, but it seems plausible that knowledge of breast cancer would impact BSE as opposed to the other way around. This supports the fact that knowledge about breast cancer can play vital role in the increased breast awareness and the practice of BSE. As Bangladesh is a developing country with low resources, mammography as a screening procedure is not a feasible first-line prevention or detection procedure. Therefore, BSE and CBE should be the main screening procedure for breast cancer in Bangladesh. Television/radio/mass media were identified as the main source of information in this present study and this was similar with other studies [15, 25, 26], which highlights the key mechanisms for informing women about breast cancer risks, prevention and treatment, and also the importance of BSE.

The number of years that our participants had been studying had a significant association (*p*<.001) with breast cancer knowledge about symptoms, risk factors and treatment options indicating that the higher the educational level is associated with higher the knowledge about breast cancer among respondents. In addition, those who had a family history of breast cancer with close relatives had significantly higher knowledge about BSE procedure (*p*<.001). A similar association was also found in Saudi Arabia [27].

A Malaysian study found that knowledge of breast cancer was low among young and less educated women [28]. On the contrary, a study conducted in India found that as no significant association found between demographic variables and level of knowledge of breast cancer and BSE among women [29], and younger women in college [30]. These indicate that in different populations, the association level varies for many reasons like the characteristics of socio-demography and also may be other socio-cultural causes. The present study indicated that the knowledge gap was the main driving factors that impeded the BSE practice among respondents of this study, although we only included women enrolled at university so we cannot comment on educational and socio-economic differences with other women in Bangladesh.

### Limitations

All the data were self-reported by the respondents and there is chance of recall bias. No verification could be undertaken to validate respondent’s claims to practice (or not) BSE. Also, the accuracy, timing, frequency and interpretation of BSE practice were not assessed. Moreover, we conducted the study on female students of only one university using cross-sectional methods. Given that our sample are highly educated, we also cannot claim any generalizability to other groups of women in Bangladesh. So, a future study with larger sample sizes, different universities and prospective methods is warranted to overcome the potential study limitations.

## Conclusions

The present study reveals that university female students had limited knowledge on breast cancer, ranging from 30% about treatment options to 37% about symptoms, with overall correct rate of knowledge being 34.3%. Moreover, the present study revealed that the practice BSE was low in our sample. This study also found a significant association between knowledge of breast cancer and practice of BSE, implying appropriate knowledge and awareness about breast cancer can lead to practice of breast cancer screening and thus early diagnosis which in terms can help to reduce the morbidity and mortality related to breast cancer. More studies need to be conducted to find out the baseline knowledge and practice about breast cancer across the general population and between different socio-demographic groups so that effective preventive policies can be developed and implemented. Also, educational interventions programs that are socially and culturally appropriate and suitable to specific age groups should be promoted and implemented to raise awareness regarding breast cancer and enhance BSE practice among all females in Bangladesh.

## Data Availability

All data produced in the present study are available upon reasonable request to the authors.

## Acknowledgments

The authors would like to express the most profound gratitude to all of the respondents who participated in this study.

## Supporting information

**S1 File. Questionnaire**.

**S2 File. Distributions (frequencies and percentages) of all knowledge related questions**.

## References

1. Hortobagyi, G.N., et al., The global breast cancer burden: variations in epidemiology and survival. Clinical breast cancer, 2005. 6(5): p. 391–401.

2. International Research On Cancer. Cancer Fact Sheet,2020. 2020; Available from: https://gco.iarc.fr/today/data/factsheets/populations/50-bangladesh-fact-sheets.pdf.

3. DeSantis, C., et al., Breast cancer statistics, 2013. CA: a cancer journal for clinicians, 2014. 64(1): p. 52–62.

4. Bray, F., et al., Global cancer statistics 2018: GLOBOCAN estimates of incidence and mortality worldwide for 36 cancers in 185 countries. CA: a cancer journal for clinicians, 2018. 68(6): p. 394–424.

5. Youlden, D.R., et al., The descriptive epidemiology of female breast cancer: an international comparison of screening, incidence, survival and mortality. Cancer epidemiology, 2012. 36(3): p. 237–248.

6. Hossain, M.S., S. Ferdous, and H.E. Karim-Kos, Breast cancer in South Asia: a Bangladeshi perspective. Cancer Epidemiology, 2014. 38(5): p. 465–470.

7. Huguley Jr, C.M., et al., Breast self-examination and survival from breast cancer. Cancer, 1988. 62(7): p. 1389–1396.

8. Anderson, B.O., et al., Early detection of breast cancer in countries with limited resources. The breast journal, 2003. 9: p. S51–S59.

9. Hussain, S.A. and R. Sullivan, Cancer control in Bangladesh. Japanese journal of clinical oncology, 2013. 43(12): p. 1159–1169.

10. Yip, C.H. and B.O. Anderson, The Breast Health Global Initiative: clinical practice guidelines for management of breast cancer in low-and middle-income countries. Expert review of anticancer therapy, 2007. 7(8): p. 1095–1104.

11. Mia, M.S., Knowledge, attitude and practice regarding breast cancer among medical students of Bangladesh: a protocol study. 2007, Centre for the Public Health.

12. Ahmed, M.S., et al., Knowledge and Practices on Breast Cancer among Bangladeshi Female University Students: A Cross-sectional Study. Asian Pacific Journal of Cancer Care, 2020. 5(1): p. 19–25.

13. Tithi, N.S., et al., A Cross-sectional Survey on Knowledge regarding Breast Cancer and Breast Self-examination among Bangladeshi Women. Breast cancer, 2018. 236(26.17): p. 22.45.

14. Nemenqani, D.M., et al., Knowledge, attitude and practice of breast self examination and breast cancer among female medical students in Taif, Saudi Arabia. Open Journal of Preventive Medicine, 2014. 2014.

15. Boulos, D.N. and R.R. Ghali, Awareness of breast cancer among female students at Ain Shams University, Egypt. Global journal of health science, 2014. 6(1): p. 154.

16. Gilani, S.I., et al., Knowledge, attitude and practice of a Pakistani female cohort towards breast cancer. JPMA. The Journal of the Pakistan Medical Association, 2010. 60(3): p. 205.

17. Karayurt, Ö., D. Özmen, and A.Ç. Çetinkaya, Awareness of breast cancer risk factors and practice of breast self examination among high school students in Turkey. BMC public health, 2008. 8(1): p. 1–8.

18. Al-Dubai, S.A.R., et al., Exploration of barriers to breast-self examination among urban women in Shah Alam, Malaysia: a cross sectional study. Asian Pacific Journal of Cancer Prevention, 2012. 13(4): p. 1627–1632.

19. Alwan, N., et al., Knowledge, attitude and practice regarding breast cancer and breast self-examination among a sample of the educated population in Iraq. EMHJ-Eastern Mediterranean Health Journal, 18 (4), 337–345, 2012, 2012.

20. Rafique, S., Z. Waseem, and F. Sheerin, Breast cancer awareness, attitude and screening practices among university students: intervention needed. Biomed J Sci Tech Res, 2018. 4(5): p. 4–7.

21. Islam, R.M., et al., Awareness of breast cancer and barriers to breast screening uptake in Bangladesh: A population based survey. Maturitas, 2016. 84: p. 68–74.

22. Sama, C.-B., et al., Awareness of breast cancer and breast self-examination among female undergraduate students in a higher teachers training college in Cameroon. Pan African Medical Journal, 2017. 28(1): p. 164–164.

23. Ahmed, B.a.A., Awareness and practice of breast cancer and breast-self examination among university students in Yemen. Asian Pacific journal of cancer prevention: APJCP, 2010. 11(1): p. 101–105.

24. Wardle, J., et al., Breast self-examination: attitudes and practices among young women in Europe. European journal of cancer prevention: the official journal of the European Cancer Prevention Organisation (ECP), 1995. 4(1): p. 61–68.

25. Iheanacho, P., A. Ndu, and A.D. Emenike, Awareness of breast cancer risk factors and practice of breast self examination among female undergraduates in university of Nigeria Enugu campus. 2013.

26. Yan, Y.Y., Breast cancer: knowledge and perceptions of Chinese women in Hong Kong. Global journal of health science, 2009. 1(2): p. 97.

27. Alam, A.A., Knowledge of breast cancer and its risk and protective factors among women in Riyadh. Annals of Saudi medicine, 2006. 26(4): p. 272–277.

28. Al-Dubai, S., et al., Awareness and knowledge of breast cancer and mammography among a group of Malaysian women in Shah Alam. Asian Pac J Cancer Prev, 2011. 12(10): p. 2531–2538.

29. Aruna, S., A study to assess knowledge regarding breast cancer and BSE among working women in Chennai. Prism’s Nurs Pract, 2010. 5: p. 34–6.

30. Shalini, D.V. and M. Nayak, Awareness and impact of education on breast self examination among college going girls. Indian journal of palliative care, 2011. 17(2): p. 150.

31. Akhtari-Zavare, M., et al., Barriers to breast self examination practice among Malaysian female students: a cross sectional study. SpringerPlus, 2015. 4(1): p. 1–6.

32. Parsa, P. and M. Kandiah, Breast cancer knowledge, perception and breast self-examination practices among Iranian women. Int Med J, 2005. 4(2): p. 17–24.

