## supplementary file 1 for "Knowledge towards breast cancer, and breast self-examination practices and its barriers among university female students in Bangladesh: Findings from a cross-sectional study"

**Table 1. Distribution of knowledges about symptoms of breast cancer**

| Variables | n | (%) |
|---|---|---|
| **Sagging of breast** | | |
| Yes | 102 | (25.5) |
| No | 121 | (30.3) |
| Don't know | 177 | (44.3) |
| **Nipple discharge other than breast milk including blood or pus** | | |
| Yes | 119 | (29.8) |
| No | 73 | (18.3) |
| Don't know | 208 | (52.0) |
| **Swelling of part of the breast** | | |
| Yes | 136 | (34.0) |
| No | 69 | (17.3) |
| Don't know | 195 | (48.8) |
| **Shrinking of breast skin** | | |
| Yes | 132 | (33.0) |
| No | 84 | (21.0) |
| Don't know | 184 | (46.0) |
| **Changes in breast shape and size** | | |
| Yes | 161 | (40.3) |
| No | 47 | (11.8) |
| Don't know | 192 | (48.0) |
| **Color change of breast including redness or flaky skin** | | |
| Yes | 193 | (48.3) |
| No | 33 | (8.3) |
| Don't know | 174 | (43.5) |
| **New lump in the breast or armpit** | | |
| Yes | 171 | (42.8) |
| No | 25 | (6.3) |
| Don't know | 204 | (51.0) |
| **Abnormal pain in breast** | | |
| Yes | 162 | (40.5) |
| No | 49 | (12.3) |
| Don't know | 189 | (47.3) |

**Table 2. Distribution of knowledges about risk factors of breast cancer.**

| Variables | n | (%) |
|---|---|---|
| **Not feeding breast milk to children** | | |
| Yes | 83 | (20.8) |
| No | 92 | (23.0) |
| Don't know | 225 | (56.3) |
| **Not being physically active** | | |
| Yes | 173 | (43.3) |
| No | 84 | (21.0) |
| Don't know | 143 | (35.8) |
| **Previous treatment with hormones or radiation** | | |
| Yes | 117 | (29.3) |
| No | 63 | (15.8) |
| Don't know | 220 | (55.0) |
| **Food habit** | | |
| Yes | 143 | (35.8) |
| No | 57 | (14.2) |
| Don't know | 200 | (50.0) |
| **Cyst in Breast** | | |
| Yes | 122 | (30.5) |
| No | 69 | (17.3) |
| Don't know | 209 | (52.3) |
| **Genetic reasons/Family history** | | |
| Yes | 117 | (29.3) |
| No | 68 | (17.0) |
| Don't know | 215 | (53.8) |
| **Alcohol consumption** | | |
| Yes | 145 | (36.3) |
| No | 60 | (15.0) |
| Don't know | 195 | (48.8) |
| **Ageing/Getting older** | | |
| Yes | 144 | (36.0) |
| No | 59 | (14.8) |
| Don't know | 197 | (49.3) |
| **Consuming birth control pill regularly** | | |
| Yes | 147 | (36.8) |
| No | 44 | (11.0) |
| Don't know | 209 | (52.3) |
| **Obesity** | | |
| Yes | 149 | (37.3) |
| No | 10 | (2.5) |
| Don't know | 241 | (60.3) |

**Table 3. Distribution of knowledges about treatment of breast cancer.**

| Variables | n | (%) |
|---|---|---|
| **Breast cancer is curable if detected at early stage** | | |
| Yes | 144 | (36.0) |
| No | 48 | (12.0) |
| Don't know | 208 | (52.0) |
| **Chemotherapy is an effective treatment of breast cancer** | | |
| Yes | 161 | (40.3) |
| No | 58 | (14.5) |
| Don't know | 181 | (45.3) |
| **Surgery is an effective treatment of breast cancer** | | |
| Yes | 149 | (37.3) |
| No | 53 | (13.3) |
| Don't know | 198 | (49.5) |
| **Hormonal therapy is an effective treatment of breast cancer** | | |
| Yes | 111 | (27.8) |
| No | 37 | (9.3) |
| Don't know | 252 | (63.0) |
| **Curable by Alternative medicines** | | |
| Yes | 69 | (17.3) |
| No | 68 | (17.0) |
| Don't know | 263 | (65.8) |
| **Curable by herbal treatment** | | |
| Yes | 86 | (21.5) |
| No | 82 | (20.5) |
| Don't know | 232 | (58.0) |

**Table 4. Distribution of knowledges about prevention of breast cancer.**

| Variables | n | (%) |
|---|---|---|
| **Breast cancer is 100% preventable** | | |
| Yes | 99 | (24.8) |
| No | 128 | (32.0) |
| Don't know | 173 | (43.3) |
| **Feeding breastmilk to child regularly** | | |
| Yes | 134 | (33.5) |
| No | 58 | (14.5) |
| Don't know | 208 | (52.0) |
| **Not wearing underwear all the time** | | |
| Yes | 93 | (23.3) |
| No | 47 | (11.8) |
| Don't know | 260 | (65.0) |
| **Early detection by BSE and Clinical examination** | | |
| Yes | 84 | (21.0) |
| No | 23 | (5.8) |
| Don't know | 293 | (73.3) |
| **Early detection and seeking medical assistance if any symptoms is found** | | |
| Yes | 288 | (72.0) |
| No | 19 | (4.8) |
| Don't know | 93 | (23.3) |
| **Maintaining ideal body weight** | | |
| Yes | 156 | (39.0) |
| No | 68 | (17.0) |
| Don't know | 176 | (44.0) |
| **Being physically active** | | |
| Yes | 241 | (60.3) |
| No | 30 | (7.5) |
| Don't know | 129 | (32.3) |
| **Vaccine** | | |
| Yes | 5 | (1.3) |
| No | 200 | (50.0) |
| Don't know | 195 | (48.8) |
| **Food habit** | | |
| Yes | 204 | (51.0) |
| No | 47 | (11.8) |
| Don't know | 149 | (37.3) |

**Table 5. Distribution of knowledge about screening of breast cancer.**

| Variables | n | (%) |
|---|---|---|
| **Clinical examination is a type of screening** | | |
| Yes | 109 | (27.3) |
| No | 125 | (31.3) |
| Don't know | 166 | (41.5) |
| **Mammography is a type of screening** | | |
| Yes | 160 | (40.0) |
| No | 65 | (16.3) |
| Don't know | 175 | (43.8) |
| **Breast self-examination is a type of screening** | | |
| Yes | 168 | (42.0) |
| No | 42 | (10.5) |
| Don't know | 190 | (47.5) |
| **Ultrasound is a type of screening** | | |
| Yes | 134 | (33.5) |
| No | 67 | (16.8) |
| Don't know | 199 | (49.8) |
| **Biopsy is a type of screening** | | |
| Yes | 156 | (39.0) |
| No | 46 | (11.5) |
| Don't know | 198 | (49.5) |

**Table 6. Distribution of knowledge about process of breast self-examination.**

| Variables | n | (%) |
|---|---|---|
| **Inspecting breast visually Infront of a mirror to look for any changes like size, shape, color, unusual discharge or nipple inversion** | | |
| Yes | 127 | (31.8) |
| No | 139 | (34.8) |
| Don't know | 134 | (33.5) |
| **Inspecting breast and look for changes Infront of mirror by holding arms at sides, by arms over head, by hands on hips and tighten chest muscle, and by bending forward with hands on hips** | | |
| Yes | 128 | (32.0) |
| No | 144 | (36.0) |
| Don't know | 128 | (32.0) |
| **Inspecting breast by lying down on back with pillow under shoulder and use pads of three middle fingers to give pressure in circle, up and down pattern for each breast** | | |
| Yes | 126 | (31.5) |
| No | 119 | (29.8) |
| Don't know | 155 | (38.8) |
| **Feel for changes in armpits by fingers in up down vertical** | | |
| Yes | 123 | (30.8) |
| No | 118 | (29.5) |
| Don't know | 159 | (39.8) |
| **Inspecting breasts while bathing with soap** | | |
| Yes | 125 | (31.3) |
| No | 108 | (27.0) |
| Don't know | 167 | (41.8) |
