## supplementary file 2 for "Knowledge towards breast cancer, and breast self-examination practices and its barriers among university female students in Bangladesh: Findings from a cross-sectional study"

**Effectiveness of Educational Intervention on Knowledge on Breast Cancer and Breast Self-Examination among Female Students in a Selected University of Bangladesh**

### Informed consent form

I am a graduate student of Dept. of Public Health and Informatics at Jahangirnagar University, Savar, Dhaka. I'm conducting a study named "**Effectiveness of Educational Intervention on Knowledge on Breast Cancer and Breast Self-Examination among Female Students in a Selected University of Bangladesh**" for the partial fulfillment of my M.Sc. Degree. As my thesis requirement, I am conducting this educational interventional study in three phases. For the pre-test you have to fulfill a questionnaire on breast cancer after that an educational session will be provided using slide presentation, video demonstration, and discussion and lastly a leaflet will be given to you where stepwise breast self- examination procedure will be described. For the pre-test and educational session phase, 60-70 minutes' time will be taken from you. And after 15 days of interval, you will again be asked to join the post-test phase where the same questionnaire as pre-test will be given to you and you will have to fulfill that. During post-test phase, 10-15 minutes will be needed to fill up the questionnaire. I invite you to join this study and I assure you that your responses will be confidential (subjects' identities and responses will be known to investigator but will not be divulged) or that anonymity will be (subjects' identities will not be known or connected to responses) preserved and that your name will not be associated with any results of this study. Also, you will receive no remuneration or any monetary benefit from this study except for the educational session where you will come to know about some interesting and important information on breast cancer. Your voluntary participation for this study is much appreciated. You can leave this study at any moment according to your convenient.

### Participant’s consent

The general nature of this study entitled "**Effectiveness of educational intervention on breast cancer knowledge and breast self-examination among female students in a selected University of Bangladesh**" has been explained to me. I understand that this study is an interventional study which requires my full participation in the pre-test, educational session and post-test periods. I will be asked to give answers regarding to breast cancer. My participation in this study should take a total of about 60-70 minutes for pre-test and educational session period and afterwards for the post-test it will take few minutes more to fulfill the questionnaire form. I understand that my responses will be confidential (subjects’ identities and responses will be known to investigator but will not be divulged) or that anonymity will be (subjects’ identities will not be known or connected to responses) preserved and that my name will not be associated with any results of this study. I know that I may refuse to answer any question asked and that I may discontinue participation at any time. I also understand that it is a non-paid participation from my end. Potential risks resulting from my participation in this project have been described to me. I am aware that I must be at least 18 years of age to participate. My signature below signifies my voluntary participation in this project, and that I have received a copy of this consent form.

Date ______________________________ ______________________________ Signature

**A. Socio-demographic characteristics of Respondents**

**Name:**

**Phone Number:**

**Name of the residing residential hall:**

**Hall room No:**

**Age:**

**Dept.:** **University study year:**

**Marital status:** **Children:**

**Monthly family income:** **Religion:**

**B: Knowledge about breast cancer:**

**1. Have you ever heard of breast cancer?**

a. Yes b. no

**2. Sources of information**

1 = friends/relatives 2 = social media

3 = television/radio/print media 4 = doctors

5= others

**3. Have anyone from your family suffered from breast cancer?**

a. yes b. no

**4. What is her relationship with the affected person?**

--

**5. Do you know the symptoms of breast cancer?**

a. yes b. no

**6. Knowledge about Symptoms of breast cancer:**

| Questions Symptoms | Coding/answer categories | | |
|---|---|---|---|
| Sagging of breast | 1=yes | 2=no | 3=don’t know |
| Nipple discharge other than breast milk including blood or pus | 1 = yes | 2= no | 3= don’t know |
| Swelling of part of breast | 1 = yes | 2=no | 3= don’t know |
| Shrinking of breast skin | 1 = yes | 2=no | 3= don’t know |
| Changes in breast shape and size including inverted nipple | 1=yes | 2=no | 3=don’t know |
| Color change of breast including redness or flaky skin | 1=yes | 2=no | 3=don’t know |
| New lump in the breast or armpit | 1=yes | 2=no | 3=don’t know |
| Abnormal pain in one part of breast | 1 = yes | 2=no | 3=don’t know |

**7. Knowledge about Risk factors of breast cancer:**

| | | | |
|---|---|---|---|
| Not feeding breast milk to child | 1=yes | 2=no | 3=don’t know |
| Not being physically active | 1=yes | 2=no | 3=don’t know |
| Previous treatment with hormones or radiation | 1=yes | 2=no | 3=don’t know |
| Food habit | 1=yes | 2=no | 3=don’t know |
| Cyst in breast | 1=yes | 2=no | 3=don’t know |
| Genetic reasons/ family history | 1=yes | 2=no | 3=don’t know |
| Alcohol consumption | 1=yes | 2=no | 3=don’t know |
| Ageing/getting older | 1=yes | 2=no | 3=don’t know |
| Consuming birth control pill regularly | 1=yes | 2=no | 3=don’t know |

| | | | |
|---|---|---|---|
| Obesity | 1=yes | 2=no | 3=don't know |

**8. Knowledge about Treatment of breast cancer:**

| | | | |
|---|---|---|---|
| Early detection is the best approach for breast cancer control | 1=yes | 2=no | 3=don't know |
| Chemotherapy is an effective treatment of breast cancer | 1=yes | 2=no | 3=don't know |
| Surgery is an effective treatment of breast cancer | 1=yes | 2=no | 3=don't know |
| Hormonal therapy is an effective treatment of breast cancer | 1=yes | 2=no | 3= don't know |
| Curable by Alternative medicines | 1=yes | 2=no | 3= don't know |
| Curable by Herbal medicine | 1=yes | 2=no | 3= don't know |

**9. Knowledge about breast cancer prevention:**

| | | | |
|---|---|---|---|
| Breast cancer is 100% preventable | 1=yes | 2=no | 3=don't know |
| Feeding breast milk to child regularly | 1=yes | 2=no | 3=don't know |
| Early detection by BSE and clinical examination | 1=yes | 2=no | 3=don't know |
| Early detection and seeking medical assistance if any symptoms are found | 1=yes | 2=no | 3=don't know |
| Maintaining ideal body weight | 1=yes | 2=no | 3=don't know |

| | | | |
|---|---|---|---|
| Being physically active | 1=yes | 2=no | 3=don't know |
| Vaccine | 1=yes | 2=no | 3=don't know |
| Healthy Food habit | 1=yes | 2=no | 3=don't know |

**C. Knowledge and practice about breast cancer Screening:**

**10. Do you know about screening for breast cancer?**

a. yes

b. no

c. don't know

**11. Types of breast cancer examination:**

| | | | |
|---|---|---|---|
| Clinical examination | 1=yes | 2=no | 3=don't know |
| Mammography | 1=yes | 2=no | 3=don't know |
| Breast self-examination | 1=yes | 2=no | 3=don't know |
| Breast ultrasound | 1=yes | 2=no | 3=don't know |
| MRI | 1=yes | 2=no | 3=don't know |

**12. Have you ever self-examined your breast for breast cancer?**

**a. Yes**

**b. No**

**13. How breast self-examination is done?**

| | | | |
|---|---|---|---|
| Inspecting breast visually In front of a mirror to look for any changes like size, shape, color, unusual discharge or nipple inversion | 1=yes | 2=n0 | 3=don't know |
| Inspecting breast and look for changes In front of mirror by holding arms at sides, by arms over head, by hands on hips and tighten chest muscle, and by bending forward with hands on hips | 1=yes | 2=no | 3=don't know |

| | | | |
|---|---|---|---|
| Inspecting breast by lying down on back with pillow under shoulder and use pads of three middle fingers and hand palm to give pressure in circle, up and down pattern for each breast and look for lump | 1=yes | 2=no | 3=don’t know |
| Feel for changes/lump in armpits by hand palm and three middle finger pads in up down vertical motion | 1=yes | 2=no | 3=don’t know |
| Inspecting breasts while bathing with soap | 1=yes | 2=no | 3=don’t know |

**14. Barrier of Breast self-examination practice:**

a. I don’t have any symptom
b. Lack of privacy/ shyness/ uncomfortable feelings
c. Lack of knowledge about the screening
d. Fear or being diagnosed of cancer
e. I don't think it's important
f. I know i will never have breast cancer
g. Other
